# Impact of Genetic Risk Factors on Coronary Heart Disease Risk Across the Age Spectrum in Three Major Race/Ethnicity Groups in the United States

**DOI:** 10.1101/2025.11.10.25339939

**Authors:** Mohammadreza Naderian, Johanna L. Smith, Marwan E. Hamed, Ozan Dikilitas, Josh B. Cortopassi, Elizabeth M. McNally, Qiping Feng, Ryan Irvin, Gail P. Jarvik, Leah C. Kottyan, Nita A. Limdi, Emily Miller, Bahram Namjou-Khales, Megan Roy-Puckelwartz, Robb Rowley, Sarah C. Hanks, Eimear E. Kenny, Noura S. Abul-Husn, Hemant K. Tiwari, Wei-Qi Wei, Atlas Khan, John J. Connolly, Georgia Wiesner, Teri A. Manolio, Richard R. Sharp, Iftikhar J. Kullo

## Abstract

**Background:** Measures of genetic predisposition can improve prediction of risk of cardiometabolic diseases but more data is needed in groups under-represented in genomics research. In this study, we investigated the impact of genetic risk factors for coronary heart disease (CHD) - polygenic risk, monogenic risk [in the form of familial hypercholesterolemia (FH)], and family history (FamHx) - on CHD risk estimates, across the age spectrum, in two diverse cohorts of US adults - eMERGE IV (eIV) and *All of Us* (AoU).

**Methods:** CHD was defined as myocardial infarction, unstable angina, and coronary revascularization. Self-identified race/ethnicity (SIRE) was used as a population descriptor. We calculated a polygenic risk score for CHD (PRS_CHD_, PGS004698), ascertained FH as presence of pathogenic/likely pathogenic variants in FH genes, and defined FamHx as early-onset CHD in a first-degree family member. We employed Pooled Cohort Equations (PCE) to estimate the 10-year risk of CHD for adults ≥40 y and modeled the association of conventional risk factors with CHD in adults <40 y. We analyzed the impact of PRS_CHD_ and FamHx on CHD risk estimates by a) using multivariable logistic regression and Cox proportional hazard models, assessing discrimination and the extent of risk reclassification; and b) net benefit analysis and decision curves to assess the performance of prediction models across actionable thresholds.

**Results:** We analyzed data for 19,348 participants from eIV (age 50.6±15.0, 68% female, 40.5% non-White) and 239,645 participants from AoU (age 55.4±17.0, 60.6% female, 48% non-White). The effects of PRS_CHD_ and FamHx on CHD were independent and additive in the two cohorts and incorporating both into PCE for eIV participants significantly improved discrimination (C-statistic increased from 0.719 to 0.753; *P*-diff=9.1×10^−3^) and reclassified risk in 18.7% and 20.2% of participants at the 7.5% and 10% 10-y CHD risk thresholds, respectively. Between the 7.5% and 10% 10-y CHD risk thresholds, incorporating PRS_CHD_ and FamHx into the PCE improved the net benefit of the risk prediction models across all SIRE groups.

**Conclusion:** PRS_CHD_ and FamHx were independently and additively associated with CHD across major SIRE groups in two diverse cohorts in the US. Incorporating PRS_CHD_ and FamHx into PCE improved risk discrimination, reclassified risk in a significant portion of participants at actionable 10-y CHD risk thresholds, and improved net benefit of the PCE, motivating the addition of these factors to clinical risk algorithms.

## Introduction

The prevalence of cardiometabolic diseases such as obesity, diabetes, and coronary heart disease (CHD) continues to remain high in the United States and many other countries around the world.(1) Early detection of individuals at increased risk can enable targeted preventive measures to reduce the burden of such diseases. However, prediction of cardiometabolic disease risk has modest accuracy due to lack of sufficiently predictive biomarkers. One approach to improving risk prediction is to incorporate measures of genetic predisposition into algorithms to predict risk of cardiometabolic diseases.(2) For example, the cumulative effects of numerous single-nucleotide variants (SNVs) with modest effects, summated as a polygenic risk score (PRS), can provide incremental predictive utility for such diseases.(3) Additional measures of genetic predisposition include family history (FamHx), which reflects shared environment or unmeasured genetic components and their interactions, and rare pathogenic/likely pathogenic (P/LP) variants.

Over the last several years, there has been increasing interest in the clinical use of genetic risk markers such as PRSs (4,5) and health systems and the private sector have begun to incorporate PRSs into disease prevention programs.(6,7) A major factor hindering their routine use is the unclear predictive utility of both PRS and FamHx in diverse cohorts. Most evidence is derived from cohorts comprising predominantly of participants of European genetic ancestry, limiting generalizability to other genetic ancestry and self-identified race/ethnicity (SIRE) groups. Therefore, it remains unclear how the three genetic risk factors—PRS, monogenic etiology, and FamHx—influence cardiometabolic risk prediction across different SIRE groups.

To investigate the predictive utility of integrating genetic risk factors into risk algorithms across SIRE groups in the US, we investigated the impact of these factors on CHD risk. Genetic prediction for CHD could be mediated through a (PRS),(8) P/LP variants in familial hypercholesterolemia (FH) genes,(9) as well as a positive FamHx.(10,11) FH is strongly associated with CHD risk and is an indication for high-intensity statin therapy per guidelines on primary prevention of cardiovascular disease.(12) On the other hand, FamHx although robustly associated with CHD risk, is classified merely as a risk enhancer,(13) and polygenic risk is absent from the guidelines, despite evidence that individuals in the top 5^th^ percentile of PRS for CHD have a risk comparable to those with FH.(14) For a comprehensive assessment of disease risk, it is important to consider PRS alongside other genetic factors.

In a prior analysis of data from phase II of the electronic Medical Records and Genomics (eMERGE) Network, we demonstrated that despite performing less well in African Americans, a PRS for CHD led to significant risk reclassification in this group.(15) In a recent analysis of the diverse eMERGE phase IV study cohort, we noted that at least one of the three genetic risk factors (PRS, FH, and FamHx) was present in ∼14% of participants (unpublished data). We analyzed data from two diverse cohorts: eMERGE phase IV study cohort, designed to assess outcomes after returning comprehensive genetic risk information for nine common diseases in adults, as well as the *All of Us* (AoU) research program cohort, designed to advance biomedical research and healthcare for underrepresented populations. Such investigation is imperative to generate the evidence base for equitable implementation of precision medicine for cardiometabolic disease including CHD.

## Methods

### Data Availability

The data, analytic methods, and study materials underlying this article are not publicly available. However, de-identified data and analytic code may be made available to qualified researchers upon reasonable request to the corresponding author and after approval by the eMERGE consortium.

### Data source

The eMERGE network was initiated in 2007 by the National Human Genome Research Institute (NHGRI), to utilize electronic health records (EHR) for genomic discovery and implementation.(16,17) In eMERGE phase IV (eIV), the network developed trans-ancestry PRSs for 9 common diseases including CHD, in adults from different SIRE groups and compiled Genome Informed Risk Assessment (GIRA) reports for each condition.(18) GIRA reports were returned to all enrolled participants, regardless of their genetic risk status. As of April 2025, 20,421 participants aged 18-75 years were recruited at 10 eMERGE clinical sites. The sample for the present study consists of all recruited participants to whom a GIRA report was returned. The study prospectively recruited individuals for genotyping and exome sequencing.(18) After consenting, participants provided a blood or saliva sample for DNA, provided detailed FamHx, completed multiple surveys, and granted access to their EHR. Data including demographics and prior diagnoses was abstracted from EHR using a customized abstraction form created in REDCap™.(19) The study protocol, informed consent form, and all surveys were reviewed and approved by independent ethics committees or institutional review boards at each site and the central Institutional Review Board approved the study before data collection. In the eIV cohort, we excluded participants with missing age, sex, SIRE, and genetic data (**Figure 1**).

**Figure 1:**
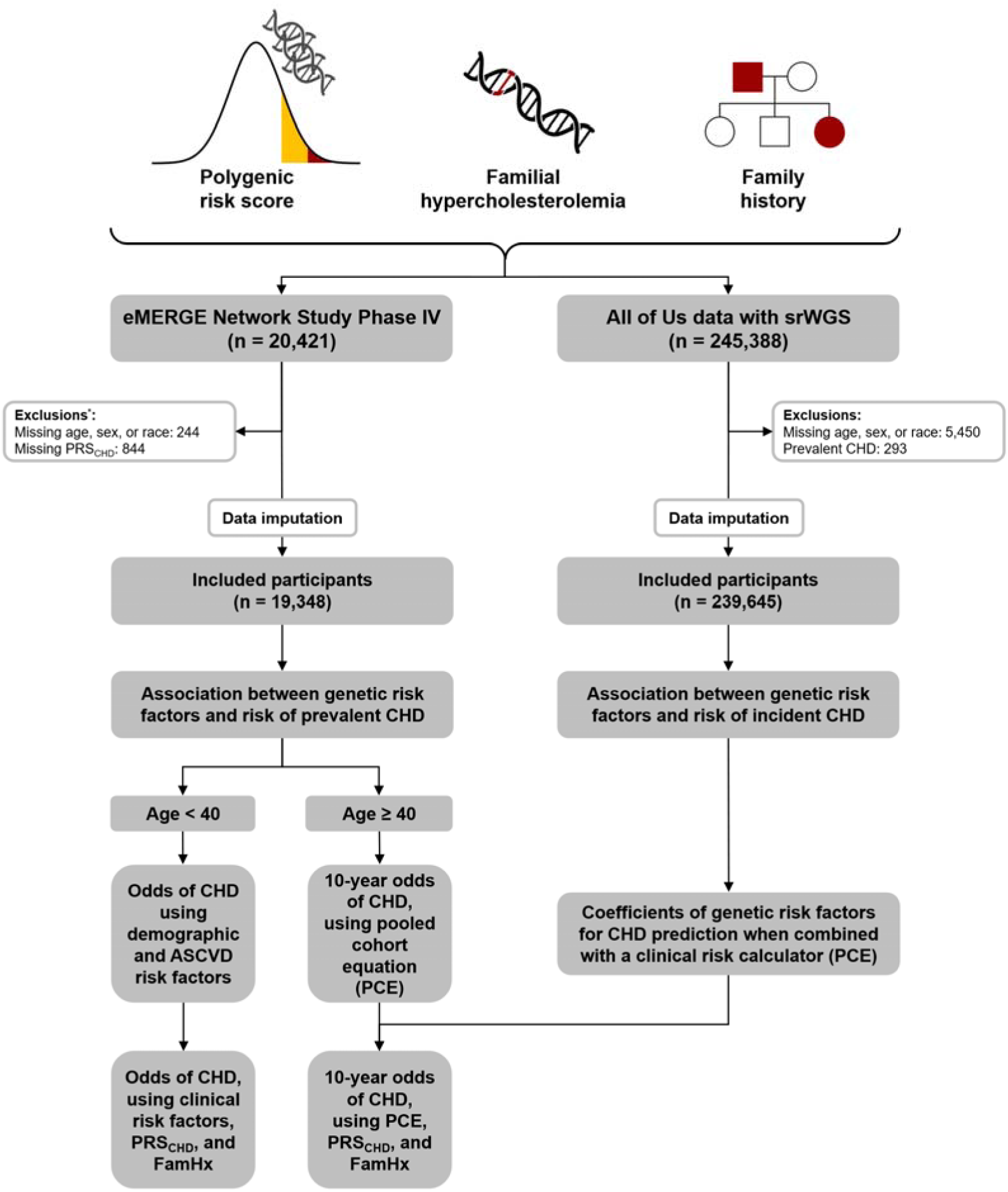
Study Design Three genetic risk factors—PRS_CHD_, FH, and FamHx—were ascertained in two diverse cohorts in the U.S.: eIV and AoU. We assessed the association of genetic risk factors with all CHD cases (prevalent CHD) in eIV, and incident CHD in AoU. We utilized AoU data to derive coefficients for PCE, PRS_CHD_, and FamHx when incorporated into a risk prediction model, PCE, for individuals aged 40 and older in eIV. **Abbreviations:** CHD: coronary heart disease, FamHx: family history, PRS_CHD_: polygenic risk score for CHD, PCE: pooled cohort equations, srWGS: short-read whole genome sequencing

The AoU Research Program aims to enroll one million diverse U.S. participants engaging with 22 community partners to collect extensive health data, prioritizing underrepresented groups, with targets of over 45% racial minorities and 75% underrepresented populations.(20,21) Researchers access surveys, medical history, physical measurements, biospecimens, and structured healthcare data.(22) Of 413,450 participants in version 7 of AoU data, we included 245,388 with whole-genome sequencing data after excluding participants with missing age, sex, or SIRE, and participants with prevalent CHD (**Figure 1**).

### Population descriptors

In both eIV study and AoU, sex was determined based on self-reported assigned sex at birth. In eIV, SIRE groups included White, Hispanic/Latino, Black, Asian, Middle-eastern, American, and Hawaiian. In AoU, SIRE groups included non-Hispanic White, Hispanic/Latino, non-Hispanic Black, Asian, Middle-eastern, Hawaiian and Others.

### Definition of CHD

CHD was defined as the composite of myocardial infarction, unstable angina, or surgical/percutaneous coronary revascularization. In eIV, CHD was ascertained using self-reported data provided by participants, whereas, in AoU, an EHR phenotyping algorithm based on the International Classification of Diseases, Tenth Revision (ICD-10), and Current Procedural Terminology (CPT) codes was used to ascertain CHD (**Supplemental Table 1**).(15) We included all CHD cases in eIV (prevalent CHD); in AoU, participants with preexisting CHD within 6 months of EHR first entry were excluded, to increase probability that only incident CHD cases were included.

### PRS for CHD

In eIV, participants provided blood or saliva samples for DNA and genotyping was performed at the Broad Institute using the Global Diversity Array (GDA), followed by imputation and calculation of PRS.(23) Imputation accuracy was improved by curating the panel and removing inaccurately genotyped sites based on gnomAD v.2. Performance was assessed by comparing imputed variant call formats (VCFs) from 20 matched blood and saliva pairs with blood-derived whole-genome data.(23) AoU Genome Centers standardized protocols for DNA extraction and sequencing, ensuring clinical accuracy and consistency. Quality control methodologies and validation experiments were established, with sequencing performed on the Illumina NovaSeq 6000. The Illumina DRAGEN pipeline was used for initial quality control analysis at multiple levels, including lane, library, flow cell, barcode, and sample.(24) The eIV and AoU studies applied an ancestry-based z-score calibration method to normalize PRSs. In summary, this approach, a modified PCA-based method, models both the mean and variance of PRSs as functions of genetic ancestry, allowing calculation of individual z-scores.

Adjusting for ancestry-dependent variance prevents inconsistent classification of high-risk individuals, particularly in admixed populations. A key benefit of this method is its ability to improve calibration within ancestries, especially for highly admixed individuals, by removing the dependence between admixture fraction and PRS. The AoU cohort was used to train the model, with an ancestry-balanced subset created using PCA and a support vector machine to assign participants to 1000 Genomes (1KG) super populations. High admixture in the training and test cohorts reflected the US population diversity, validating performance across ancestries. Using an identical imputation pipeline to eIV minimized bias, ensuring that PRSs are effectively calibrated and applicable in multi-ancestry populations. In both eIV study and AoU data, a trans-ancestry PRS for CHD (PRS_CHD_, PGS004698) adjusted for the first 5 principal components of ancestry was computed for CHD (**Supplemental Figure 1**);(25) individuals with a PRS_CHD_ in the top 5^th^ percentile for their genetic ancestry were considered as having a high PRS_CHD_.

### Monogenic Risk

The Invitae laboratory performed sequencing of FH genes (*APOB, LDLR, PCSK9,* and *LDLRAP1*),(26,27) other tier 1 condition genes and *LMNA*, identified P/LP variants, confirmed these by Sanger sequencing, and issued clinical reports for disclosure to participants and placement in the EHR. Variants of uncertain significance were not returned. In AoU data, P/LP variants in FH genes were identified using a systematic variant filtering workflow. High-impact variants, including non-missense types such as stop-gain, frameshift, and splice-site variants, were retained, due to their higher likelihood of causing loss of function. Missense variants, which may or may not disrupt protein function, were further filtered based on pathogenicity predictions using REVEL (>0.50)(28) or AlphaMissense >0.56,(29) with minor allele frequency <0.005 in either gnomAD(30) or AoU databases). All variants, regardless of type, were ultimately required to have a P/LP classification in ClinVar with assertion criteria and no conflicts (accessed January 2, 2025)(31). This approach ensured that high-impact variants were appropriately captured as putative loss-of-function alleles without the use of additional computational tools, while predicted deleterious missense variants were also considered, providing comprehensive coverage of relevant FH variants for all related genes.

### Family history

In eIV participants, family history of CHD was ascertained from the MeTree software or a backup survey.(32,33) Family history of premature CHD (FamHx), was defined as CHD in male first-degree family members aged <55 years and females aged <65 years. A pedigree was generated and included in the GIRA report. In AoU, the same definition was used to ascertain FamHx from self-reported data.

### Clinical risk prediction models for CHD

In both eIV and AoU, the Pooled Cohort Equations (PCE) were used to estimate the 10-y CHD risk in adults aged over 40 years.(34) Data required for PCE risk calculations were sourced from the EHR and supplemented by survey responses where needed. In participants younger than 40 years, we constructed prediction models that included conventional risk factors for atherosclerotic cardiovascular disease (ASCVD) – age, sex, SIRE, systolic blood pressure, total cholesterol, smoking status, anti-hypertensive therapy, and lipid-lowering therapy. Various classes of blood pressure lowering agents were used to define anti-hypertensive therapy. Lipid-lowering therapy refers to the use of statins, ezetimibe, or proprotein convertase subtilisin/kexin type 9 (PCSK9) inhibitors.

### Integration of genetic risk factors into clinical risk prediction models

Given the smaller size of eIV cohort, we used AoU as training/development cohort to develop integrated risk scores. Furthermore, as risk prediction models require prospective data to estimate risk of incident disease, we used incident CHD cases from AoU to derive weights for ancestry-specific PRS_CHD_ and FamHx when combined with PCE. We then used these obtained weights from a refitted model to incorporate PRS_CHD_ and FamHx into the PCE for eIV participants. We assessed the increment in C-statistic after incorporation of PRS_CHD_ and after incorporation of both PRS_CHD_ and FamHx (See Statistical Analyses, Assessment of Prediction Model Performance).

### Independent and additive effects of genetic risk factors

In both eIV and AoU, we assessed the independent and additive effect of PRS_CHD_ and FamHx on CHD. We assessed the association of CHD with high PRS_CHD_ using multivariable logistic regression models in eIV and multivariable Cox proportional hazards models in AoU, stratifying analyses by the presence or absence of FamHx. To examine the additive effect of PRS_CHD_ and FamHx, we used individuals without high PRS_CHD_ and without FamHx as the reference group. In both logistic regression and Cox proportional hazards models, we assessed for interactions between PRS_CHD_ and FamHx and assessed their additive effects on CHD.

### Statistical Analyses

We imputed the missing data using predictive mean matching for continuous variables and polytomous regression for ordered and unordered categorical variables.(35) Genetic risk factors were defined as PRS_CHD_, FH, or FamHx. We used multivariable logistic regression to test the association between genetic risk factors and prevalent CHD in eIV, and multivariable Cox proportional hazard model to test the association between genetic risk factors and incident CHD in AoU. Hazard ratios (HR) were reported per 10 years of follow-up.

### Assessment of Prediction Model Performance

To assess discrimination power of different prediction models, we used the C-statistic. Additionally, 95% confidence intervals (CIs) for the difference in C-statistics were computed to quantify the uncertainty around the estimated differences. To compare the C-statistic of different models, we used the DeLong test, a non-parametric approach that assesses whether the difference between two C-statistics is statistically significant.(36) We also calculated reclassification percentages using 10-y CHD risk of 7.5% and 10% as the actionable thresholds, based on the ACC/AHA guideline on primary prevention of cardiovascular disease. (12) Participants were categorized into low and high-risk groups based on each threshold, allowing us to estimate reclassification.

We also calculated net benefit, an increasingly used metric to evaluate the performance of risk prediction models.(37) Net benefit analysis quantifies the clinical utility of a prediction model by balancing the benefits of true positives against the harms of false positives. (15) Unlike traditional metrics such as sensitivity and specificity, which do not account for the consequences of clinical decisions, net benefit directly incorporates the clinical context by weighting false positives according to the threshold probability. Net benefit was calculated to evaluate the clinical utility of the prediction models at each decision threshold. Net benefit combines the true positive rate (benefit of correctly identifying high-risk individuals) and the false positive rate (harm of incorrectly labeling low-risk individuals as high-risk) into a single metric, adjusting for the threshold probability.

The net benefit for prediction models was calculated using the following formula:

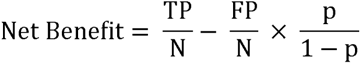

In the equation, TP is the number of true positives (participants correctly identified as high risk and who had an event), FP is the number of false positives (participants incorrectly identified as high risk but did not have any event), N is the total number of participants, and p is the actionable threshold for preventive strategies. Additionally, the net benefit for the strategies of ‘preventing all participants’ and ‘preventing none of participants’ were calculated as reference points. We used decision curve analysis (DCA) to plot the net benefit of integrated risk prediction models compared to the PCE at each decision threshold, alongside the net benefits of the ‘preventing all’ and ‘preventing none’ strategies. We emphasized the net benefit at the 10-y CHD risk of 7.5% and 10% actionable thresholds, which are commonly used in clinical decision-making for considering statin initiation.

## Results

### Participants Characteristics

After applying exclusion criteria, 19,348 participants from eIV and 239,645 participants from AoU were included in the final analyses. The mean age of the eIV participants was 50.6±15.0 years (range 18 - 75 years) and 68.0% were female. SIRE was 59.5% White, 18.2% Hispanic/Latino, 13.3% Black, 7.9% Asian, and 1.1% Middle-eastern, American, and Hawaiian. Based on self-report, 775 (4.0%) of participants had CHD. The mean age of the AoU participants was 55.4±17.0 years (range 18–118 years) and 60.6% were female. SIRE was 52.0% non-Hispanic White, 19.5% Hispanic/Latino, 20.5% non-Hispanic Black, and 8.0% as Other. In AoU, median [interquartile range] follow-up duration was 7 [3 - 10] years, during which 6,732 participants (2.8%) had incident CHD (**Supplemental Table 2**, **Supplemental Table 3).**

The estimated median [interquartile range (IQR)] of the 10-year CHD risk based on the PCE was 6.6% [2.1% - 14.7%] for eIV participants older than 40 y, and 7.5% [2.7% - 16.0%] for AoU participants.

In both cohorts, at baseline, participants with high PRS_CHD_ were younger, had a higher burden of cardiac risk factors, and were more likely to be on LLT, anti-hypertensive, or anti-diabetic medications, with no significant difference in baseline 10-year ASCVD risk (**Supplemental Table 4**). In addition, participants with FamHx were older, had a higher burden of cardiac risk factors, and were less likely to be of non-White SIRE, and more likely to be on LLT, anti-hypertensive therapy, and anti-diabetic medications. Baseline 10-year ASCVD risk was therefore significantly higher in participants with FamHx (**Supplemental Table 5**). Participants with FH were less likely to be of non-White SIRE, and more likely to be on LLT and have a higher baseline adjusted LDL-C. However, there was no significant difference in 10-year ASCVD risk ≥ 7.5 between those with and without FH (**Supplemental Table 6**).

### Association of genetic risk factors with CHD

In eIV, 2814 (14.5%) participants had at least 1 of the 3 genetic risk factors - 861 (4.5%) had high PRS_CHD_, 139 (0.7%) had P/LP variant in genes associated with FH, and 1991 (10.3%) had FamHx (**Supplemental Table 7**, top section). In a logistic regression model adjusted for age and sex, the odds ratio (OR) for 1 standard deviation (SD) increase in PRS_CHD_ with CHD was 1.39 (95% confidence interval (CI): 1.29 - 1.50, *P*<2.2×10^−16^), and for high PRS_CHD_ (top 5^th^ percentile) was 1.57 (95% CI: 1.14 – 2.13, *P*=4.2×10^−3^). For FH, the OR was 2.25 (95% CI: 1.17 – 4.00, *P*=8.8×10^−3^), and for FamHx was 2.38 (95% CI: 1.99 – 2.84, *P*<2.2×10^−16^) (**Table 1**, top section). In subgroup analyses, the association of a 1 SD increase in PRS_CHD_ with CHD was strongest in White SIRE and non-significant in Hispanic/Latino and Black SIREs. The association of FH with CHD was significant in Hispanic/Latinos, while FamHx was strongly associated with CHD in both Whites and Blacks. In a multivariable logistic regression model including all three genetic risk factors as predictors, each remained independently associated with CHD, with ORs of approximately 1.3 for PRS_CHD_, 2.0 for FH, and 2.5 for FamHx (**Supplemental Table 8**, top section).

**Table 1:**
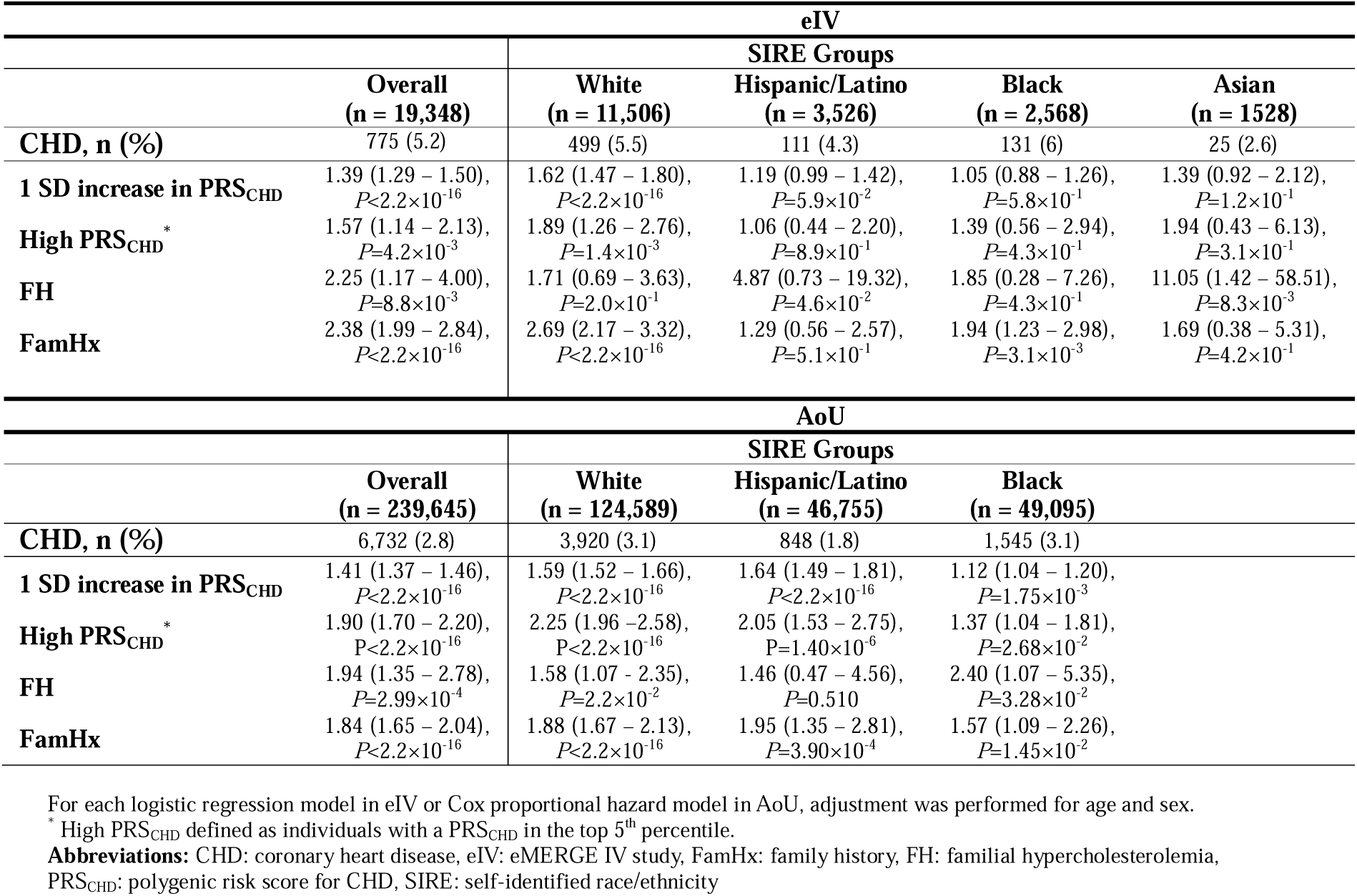
Association of Genetic Risk Factors with Coronary Heart Disease Across SIRE groups in eIV and AoU.

In AoU, 42,747 (17.8%) of participants had at least 1 of the 3 genetic risk factors: 11,983 (5.0%) had high PRS_CHD_, 953 (0.4%) had FH (**Supplemental Table 7**, bottom section) and 32,417 (13.5%) had FamHx. In a Cox proportional hazard model adjusted for age, sex, and 5 PCs, the HR for the association of 1 SD increase in PRS_CHD_ with CHD was 1.41 (95% CI: 1.37 - 1.46, *P*<2.2×10^−16^), and for high PRS_CHD_ was 1.90 (95% CI: 1.70 – 2.20, *P*<2.2×10^−16^). For FH, the HR was 1.94 (95% CI: 1.35-2.78, *P=*2.9×10^−4^), and for FamHx was 1.84 (95% CI: 1.65-2.04, *P<*2.2×10^−16^) (**Table 1**, bottom section). In subgroup analyses, a 1 SD increase in PRS_CHD_ was significantly associated with CHD in all groups, with the strongest associations observed in Whites and Hispanic/Latinos, and a weaker association in Blacks. FH was significantly associated with CHD in Whites and Blacks. FamHx was significantly associated with CHD in all groups. In a Cox proportional hazard model including all three genetic risk factors as predictors, each remained independently associated with CHD, with HRs of approximately 1.4 for PRS_CHD_, 1.9 for FH, and 1.5 for FamHx (**Supplemental Table 8**, bottom section).

### Independent and additive effects of genetic risk factors

In eIV, after adjustment for age and sex, the ORs for 1 SD increase in PRS_CHD_ and the risk of CHD were similar in participants with or without FamHx: 1.32 (95% CI: 1.21 – 1.45, *P*=3.1×10^−10^) vs. 1.46 (95% CI: 1.24 – 1.72, *P*=5.6×10^−6^), respectively (*P*-diff= 0.291). In AoU, after adjustment for age, sex, and 5 PCs, the HRs of high PRS_CHD_ for CHD were similar in participants with or without a FamHx: 2.11 (95% CI: 1.65 - 2.69, *P*=1.85×10^−9^ vs. 1.86 (95% CI: 1.42 - 2.44, *P=*6.65×10^−6^), respectively (*P*-diff= 0.498).

The additive effect of PRS_CHD_ and FamHx on CHD are shown in **Figure 2**. In eIV, compared to participants without a FamHx and without a high PRS_CHD_, those who had only high PRS_CHD_ had higher odds of CHD: 1.36 (95% CI: 0.90 – 1.99, *P*=1.3×10^−1^) followed by those who had only positive FamHx: 2.33 (95% CI: 1.93 – 2.80, *P*<2.2×10^−16^); those who had both FamHx and high PRS_CHD_ had the highest odds of CHD: 3.54 (95% CI: 2.08 – 5.76, *P*=9.9×10^−7^). In AoU, compared to participants without a FamHx and without a high PRS_CHD_, those with only positive FamHx had a HR of 1.78 (95% CI: 1.59-1.99, *P*<2.2×10^−16^), followed by those who had only high PRS_CHD_: 1.85 (95% CI: 1.41-2.43, *P*=7.66×10-6); those who had both FamHx and high PRS_CHD_ had the highest hazard of CHD: 3.79 (95% CI: 2.98-4.82, *P*<2.2×10^−16^).

**Figure 2:**
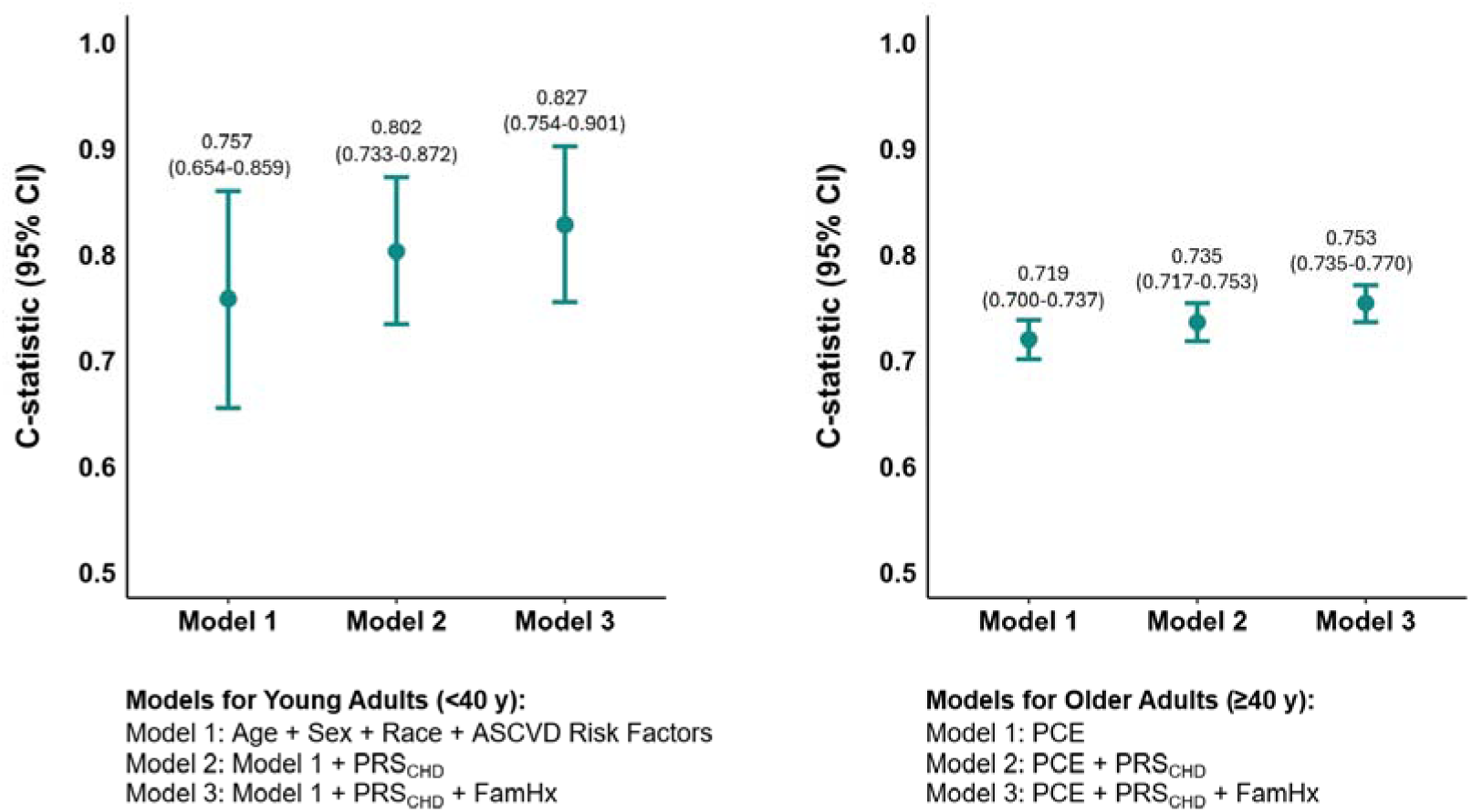
Additive Effect of PRS_CHD_ and FamHx on CHD in eIV and AoU **Abbreviations:** AoU: All of Us research program, CHD: coronary heart disease, CI: confidence interval, eIV: eMERGE IV study, FamHx: family history, PRS: polygenic risk score for CHD

### Incorporation of genetic risk factors into clinical risk estimates

In both eIV and AoU data, the correlation of PRS_CHD_ with clinical risk derived from PCE was minimal (*r*<0.05). We applied the weights of PRS_CHD_, FamHx, and PCE from the risk prediction models in AoU to validate the integrated risk scores in eIV (**Supplemental Table 9**). The distribution of risk based on the PCE and the two integrated scores is shown in (**Supplemental Figure 2**).

The effects of genetic risk factors on discrimination and risk reclassification are summarized in **Figure 3**, **Table 2**, and **Supplemental Figure 3**. The C-statistic (95% CI) of the PCE increased from 0.719 (0.700 - 0.737) to 0.735 (0.717 - 0.753) after the addition of PRS_CHD_, and to 0.753 (0.735 - 0.770) after further addition of FamHx (*P*-diff=9.1×10^−3^, **Figure 3**, right panel). Using 10-year CHD risk of 7.5% as an actionable threshold, the incorporation of PRS_CHD_ into the PCE resulted in 3.5% up-reclassification and 4.7% down-reclassification. The additional incorporation of FamHx into both PRS_CHD_ and PCE resulted in 4.6% up-reclassification and 14.1% down-reclassification. A similar analysis using 10-y CHD risk of 10% as an actionable threshold, resulted in 4.2% up-reclassification and 4.7% down-reclassification after incorporation of PRS_CHD_, and 3.2% up-reclassification and 17.0% down-reclassification after incorporation of both PRS_CHD_ and FamHx (**Supplemental Figure 3**).

**Figure 3:**
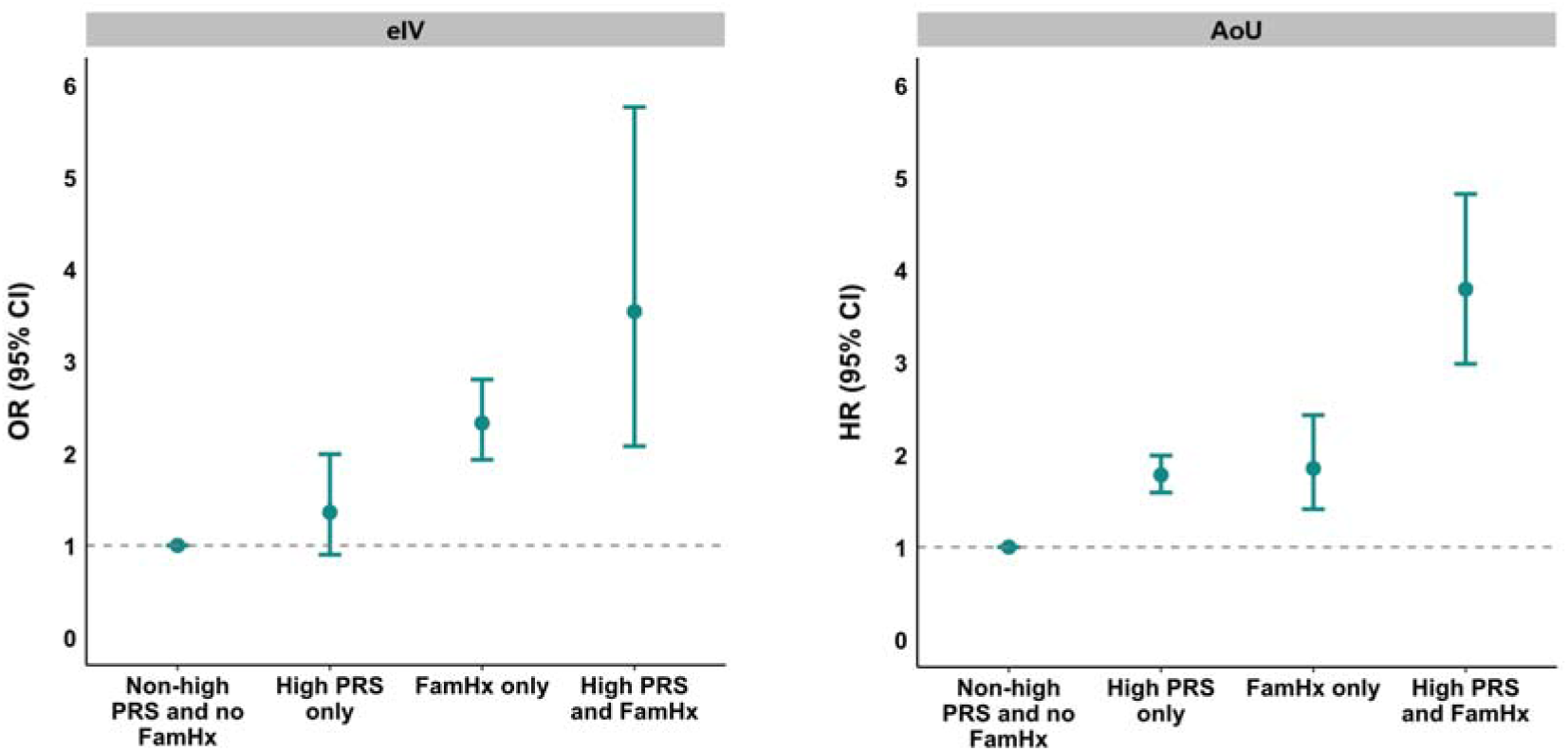
C-statistic of Logistic Regression Models for Presence of CHD in Younger and Older Adults in eIV The left panel presents the C-statistic (95% CI) for different prediction models in adults younger than 40 years. Model 1 included demographic data and ASCVD risk factors (systolic blood pressure, total cholesterol, current smoking status, and lipid-lowering therapy). Model 2 further included PRS_CHD_, and Model 3 added both PRS_CHD_ and FamHx to Model 1. The right panel shows the C-statistic (95% CI) for prediction models in adults older than 40 years. The inclusion of PRS_CHD_ (Model 2) and the combination of PRS_CHD_ and FamHx (Model 3) led to improved CHD risk discrimination. **Abbreviations:** ASCVD: atherosclerotic cardiovascular disease, CHD: coronary heart disease, CI: confidence interval, eIV: eMERGE IV study, FamHx: family history of CHD, PCE: pooled cohort equations, PRS_CHD_: polygenic risk score for CHD

**Table 2:**
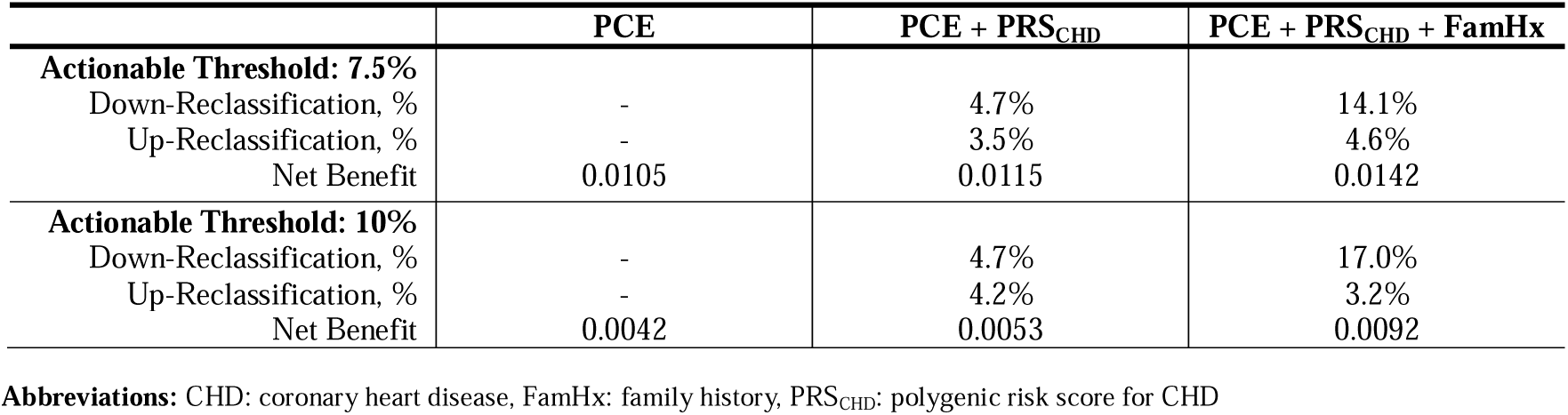
Predictive Performance of Risk Prediction Models in eIV.

|  | **PCE** | **PCE + PRS<sub>CHD</sub>** | **PCE + PRS<sub>CHD</sub> + FamHx** |
| --- | --- | --- | --- |
| **Actionable Threshold: 7.5%** |  |  |  |
| Down-Reclassification, % | - | 4.7% | 14.1% |
| Up-Reclassification, % | - | 3.5% | 4.6% |
| Net Benefit | 0.0105 | 0.0115 | 0.0142 |
| **Actionable Threshold: 10%** |  |  |  |
| Down-Reclassification, % | - | 4.7% | 17.0% |
| Up-Reclassification, % | - | 4.2% | 3.2% |
| Net Benefit | 0.0042 | 0.0053 | 0.0092 |

**Abbreviations:** CHD: coronary heart disease, FamHx: family history, PRS<sub>CHD</sub>: polygenic risk score for CHD

Results of the net benefit analysis of the PCE across actionable risk thresholds in White, Black, and Hispanic/Latino SIRE groups are shown in **Figure 4** and **Figure 5**. Incorporating PRS_CHD_ and FamHx into the PCE improved the net benefit of the integrated risk prediction models across all SIRE groups (**Figure 4**). The contribution of FamHx to net benefit improvement was greater in Hispanic/Latino and Black individuals than in Whites (**Figure 4**). Between the 7.5% and 10% CHD risk thresholds, the predictive performance of PCE was highest in Whites, followed by Blacks, with the lowest performance observed in Hispanic/Latino individuals (**Figure 5**). Similarly, the net benefit of using integrated risk scores after incorporating PRS_CHD_ or both PRS_CHD_ and FamHx into the PCE was highest in the White SIRE group, followed by Black and Hispanic/Latino SIRE groups (**Figure 5**).

**Figure 4:**
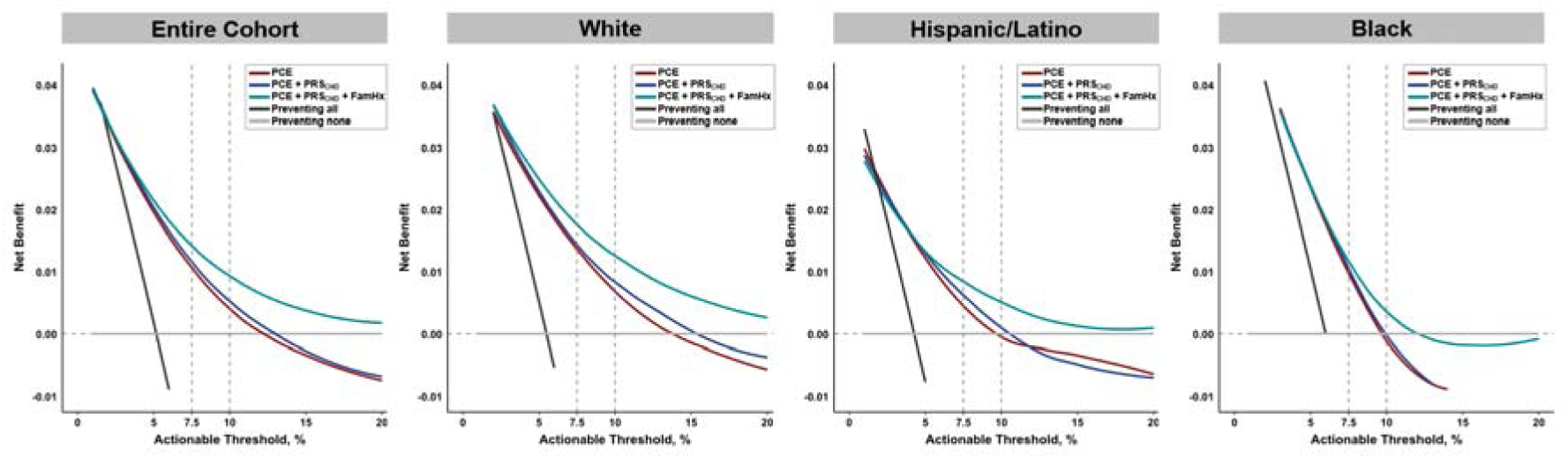
Decision Curve Analysis of CHD Prediction Models Across Actionable Risk Thresholds in eIV. This figure illustrates the net benefit of using PCE alone versus integrated models incorporating PRS_CHD_ and FamHx across actionable risk thresholds. The vertical dashed lines indicate the 7.5% and 10% actionable thresholds. The grey line represents the strategy of no intervention, yielding a net benefit of zero. The black line reflects the strategy of preventing all individuals, which initially provides the highest net benefit but declines due to unnecessary preventive interventions and associated harms or costs in low-risk persons. The red, blue, and green lines represent the net benefit of PCE, PCE + PRS_CHD_, and PCE + PRS_CHD_ + FamHx, respectively. Across the entire eIV cohort and all major SIRE groups, incorporating PRS_CHD_ and FamHx improved net benefit compared to PCE alone, highlighting the superior clinical utility of these enhanced models for guiding prevention strategies. **Abbreviations:** CHD: coronary heart disease, eIV: eMERGE IV study, FamHx: family history of CHD, PCE: pooled cohort equations, PRS_CHD_: polygenic risk score for CHD

**Figure 5:**
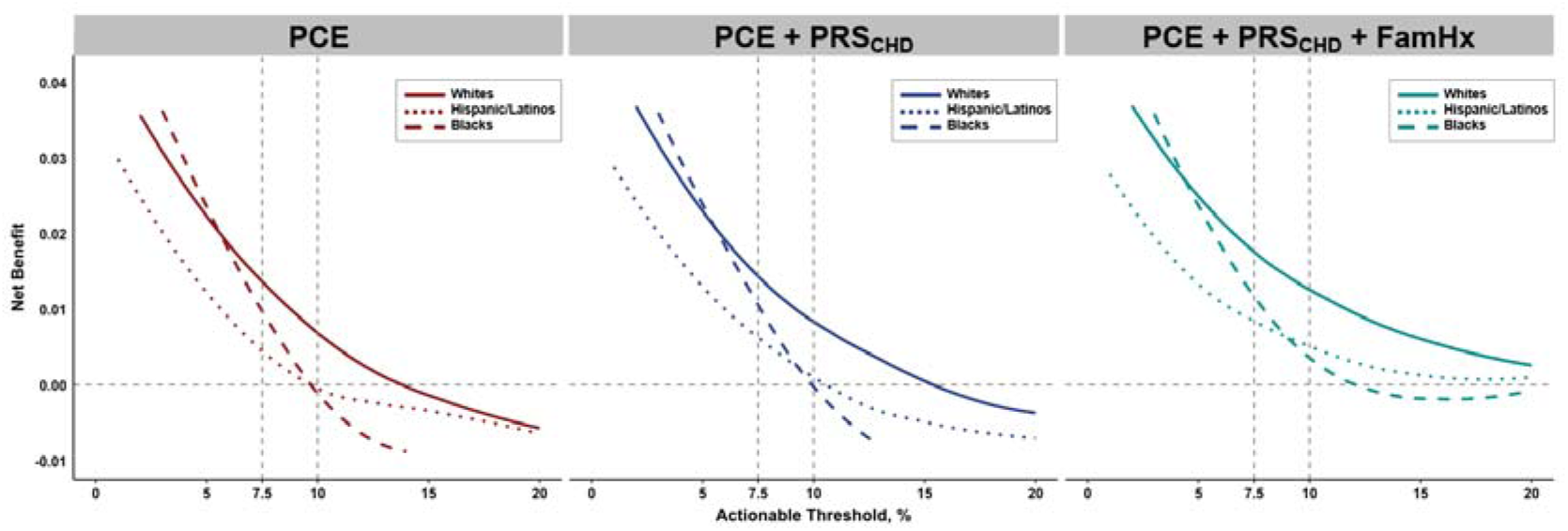
Decision Curve Analysis for PCE and Integrated Scores’ Performance Across Self-Identified Race/Ethnicity Groups in eIV. This plot presents the net benefit analysis of the PCE (left panel), PCE and PRS_CHD_ (middle panel), and PCE, PRS_CHD_, and FamHx (right panel) across actionable risk thresholds in White, Black, and Hispanic/Latino individuals. Between the 7.5% and 10% risk thresholds, PCE demonstrated the highest predictive performance in White individuals, followed by Black individuals, with the lowest performance observed in Hispanic/Latino individuals. **Abbreviation:** CHD: coronary heart disease, eIV: eMERGE IV study, FamHx: family history of CHD, PCE: pooled cohort equations, PRS_CHD_: polygenic risk score for CHD

In individuals younger than 40 years, incorporating PRS_CHD_ into a prediction model that included demographic covariates and ASCVD risk factors increased the C-statistic (95% CI) from 0.757 (0.654–0.859) to 0.802 (0.733–0.872). Further addition of FamHx increased the C-statistic to 0.827 (0.754–0.901) (*P*-diff=0.287, **Figure 3**, left panel).

## Discussion

The inclusion of PRSs, FamHx and monogenic etiology into algorithms to predict risk of cardiometabolic diseases, has great potential for realizing the promise of precision medicine at scale, but is hindered by the paucity of data in groups under-represented in genomic research.(2) We describe for the first time, the independent and additive impact of genetic risk factors on CHD risk estimates across the age spectrum in the major SIRE groups in the United States. Whereas the strength of association of PRS with CHD varied across three major SIRE groups, the association of FH and FamHx with CHD was not significantly different in the three groups. Importantly, the addition of both PRS_CHD_ and FamHx to the PCE enhanced the predictive performance of a clinical risk equation (PCE) and improved its net benefit across the three major SIRE groups.

These results extend our previous findings in the United Kingdom Biobank that PRS_CHD_ and FamHx are independently and additively associated with CHD,(9) to the major SIRE groups in the US, highlighting the potential utility of adding these factors to existing risk equations. The incorporation of PRS_CHD_ and FamHx into available clinical risk equations represents an important step toward equitable and personalized cardiovascular risk prediction, particularly in younger adults in whom conventional risk factors alone may not fully capture long-term cardiovascular risk.(14) Because of a relatively small number of individuals with FH and lack of an established approach for incorporating rare variants into a PRS or clinical risk equations, we did not include FH in our integrated risk score for CHD.(38,39)

As previously noted by us and others, a PRS for CHD performed less well in non-White groups.(15,40,41) Non-White SIRE participants had a lower prevalence of FamHx for CHD and FH in both cohorts. However, we found no significant differences in the association of FamHx and FH with CHD across SIRE groups in eIV (*P*-diff of FamHx ORs: >0.202, *P*-Diff of FH ORs: >0.06). Although the eIV cohort may have been underpowered to detect such differences, similar findings were observed in the much larger AoU cohort, where no significant differences were noted in the HRs of FamHx or FH for CHD across SIRE groups (*P*-diff of FamHx HRs: >0.400, *P*-Diff of FH HRs: >0.360). Combining PRS_CHD_ with FamHx may help reduce disparities in the performance of CHD risk calculators across SIRE groups.

The association between family history and CHD observed in the eIV cohort (OR: 2.38, (95% CI: 1.99 – 2.84), *P*<2.2×10^−16^) was stronger than in AoU (HR: 1.84, (95% CI: 1.65 – 2.04), *P*<2.2×10^−16^, **Table 1**) and the other previous population-based studies.(42,43) Adjustment for conventional cardiovascular risk factors only modestly attenuated the association, OR: 2.18 (95% CI: 1.70 – 2.80, *P*=8.5×10^−10^). This relatively strong association may have resulted from the analysis being limited to prevalent CHD and that family history was ascertained in a robust manner using the structured MeTree platform or a backup survey to capture information about premature CHD in first-degree relatives with specific age thresholds (<55 years for males, <65 years for females). When ascertained from simple questionnaires or medical record review, the true prevalence of family history as well as its strength of association with CHD may be underestimated due to incomplete or imprecise documentation. A systematic approach helps minimize recall bias and ensures standardized collection of pertinent details, including age of onset and degree of relationship.(32,44) Our findings underscore the value of family history in clinical practice to identify individuals at increased CHD risk who could benefit from preventive interventions.

The performance of the PCE for CHD risk prediction was suboptimal in Hispanic/Latino and Black groups,(45,46) as demonstrated by lower net benefit of using PCE for CHD risk discrimination in these groups than in Whites (**Figure 5**). This is not surprising since PCE were developed using data predominantly from non-Hispanic Whites and because disparities in healthcare access, baseline health conditions, and treatment responses may reduce accuracy of these models in non-Whites. Consistent with prior results in White SIRE groups, we showed in two diverse cohorts of US adults that there is weak correlation between PCE and PRS,(46) and incorporating PRS and FamHx into PCE improves the net benefit of risk prediction models compared to the PCE alone, particularly for CHD discrimination in Hispanic/Latino and Black populations (**Figure 4** and **Figure 5**). Thus, while transferability of a PRS_CHD_ across SIRE groups varies, combining it with FamHx could lessen the gap in predictive performance of clinical risk equations for CHD between SIRE groups.

The inclusion of younger adults (<40 years) in the eIV cohort enabled us to evaluate the added value of incorporating genetic risk factors in this group. We estimated 30-year CHD risk using the Framingham risk score,(10) but observed limited predictive performance in the eIV cohort (C-statistic: 0.55), possibly, because the 30-year risk model was developed in a predominantly White cohort,(47) whereas the eIV and AoU cohorts are more diverse. Additionally, the absence of cohorts with 30-year CHD incidence data restricted our ability to obtain recalibration coefficients for FRS-based risk estimates in these cohorts. While appropriate recalibration could account for differences in baseline survival across SIRE groups, these findings highlight challenges in risk communication and clinical decision-making related to longer term/lifetime CHD risk estimates in a diverse cohort. We used the recently proposed PREVENT risk equation for 10- and 30-year CHD prediction;(48) however, the results revealed poor calibration and inadequate predictive accuracy in both the eIV and AoU cohorts (analyses not shown). Nonetheless, incorporating PRS_CHD_ and FamHx into a model including demographic and ASCVD risk factors increased predictive performance, as reflected in the C-statistic (**Figure 3**), although the difference was not statistically significant, likely due to the limited number of CHD cases (n = 29) in adults <40 years in eMERGE. Further research is needed on how best to manage increased genetic risk in younger individuals.

### Strength and Limitations

Our evaluation of three genetic risk factors (polygenic risk, monogenic variants, and family history) and their incorporation into clinical risk models represents one of the most comprehensive assessments to date of genetic risk factors for CHD across major SIRE groups in the US. The complementary use of eIV and AoU allowed us to study the association of genetic risk factors with both prevalent and incident CHD. The study sample size was diverse, with ∼40% of participants self-identifying as non-White. Moreover, by obtaining relevant coefficients from the AoU data, we were able to construct integrated risk models in eIV participants. In addition, we assessed multivariable models of CHD risk in adults ≤40 years old and 10-year risk estimates in the remaining.

Several limitations should be considered when interpreting our findings. The sample size for certain SIRE groups, particularly those of Middle Eastern, American, and Hawaiian ancestry (comprising 1% of the cohort), was insufficient to draw robust conclusions about PRS performance in these populations. The use of self-reported CHD in eIV study may have introduced recall bias. However, PRS_CHD_, FH, and FamHx were independently and additively associated with incident CHD in AoU.(9) While the use of eIV dataset for validation strengthens our findings, there were differences in population risk factors and CHD ascertainment between the two cohorts. Another limitation is that eIV analyses did not include incident outcomes. Ideally, validation would be conducted in a dataset with prospectively ascertained events. However, given the unique design and diversity of ancestry in eMERGE IV, we prioritized including that dataset vs. splitting AoU into development and validation sets. Once longitudinal follow-up data in eIV becomes available, validating our findings in incident outcomes will be an important next step.

## Conclusion

Measures of genetic predisposition including PRSs, Fam Hx, and monogenic etiology, can improve risk stratification of cardiometabolic diseases, but more data is needed for groups under-represented in genomic research. The results of our study demonstrate that three genetic risk factors (PRS, FH, and FamHx) were each significantly associated with CHD in the three major SIRE groups in the US. While performance of PRS varied across SIRE groups, no significant difference was noted for the association of FamHx with CHD in these groups. Adding both FamHx and PRS to clinical risk equations (PCE) significantly improved CHD risk prediction by enhancing discrimination and reclassifying risk in a significant proportion of individuals across the three major SIRE groups. Decision curve analyses demonstrated that incorporating PRS_CHD_ and FamHx increased the net benefit of CHD prevention strategies at the 7.5% and 10% 10-y CHD risk thresholds across SIRE groups. Our findings motivate inclusion of PRS and FamHx into clinical risk calculators for equitable assessment of CHD risk.

## Data Availability Statement

The dataset generated and analyzed for the current study is available from the corresponding author on reasonable request that might be necessary to interpret, replicate, and build upon the methods or findings reported.

